# COVID-19 challenges to dentistry in the new pandemic epicenter: Brazil

**DOI:** 10.1101/2020.06.11.20128744

**Authors:** Rafael R. Moraes, Marcos B. Correa, Ana B. Queiroz, Ândrea Daneris, João P. Lopes, Tatiana Pereira-Cenci, Otávio P. D’Avila, Maximiliano S. Cenci, Giana S. Lima, Flávio F. Demarco

**Author notes:** Corresponding author: Prof. Rafael Moraes, Graduate Program in Dentistry, Federal University of Pelotas, Rua Gonçalves Chaves 457 room 505, 96015-560, Pelotas, RS, Brazil.

## Abstract

A nationwide survey of dentists was carried out in Brazil, a new pandemic epicenter, to analyze how dental coverage has been affected (public versus private networks), changes in routine and burdens, and how the local prevalence of COVID-19 affects dental professionals. Dentists were recruited via email and an Instagram® campaign. Responses to an online questionnaire were collected May 15–24, 2020. COVID-19 case/death counts in the state where respondents work was used to test associations between contextual status and decreases in weekly appointments, fear of contracting COVID-19 at work, and current work status (α=0.05). Over 10 days, 3,122 responses were received, with region, gender, and age distributions similar to those of dentists in Brazil. Work status was affected for 94% of dentists, with less developed regions being more impacted. The impact on routine was high or very high for 84%, leading to varied changes to clinic infrastructure, personal protective equipment use, patient screening, and increased costs. COVID-19 patients had been seen by 5.3% of respondents, and 90% reported fearing contracting COVID-19 at work. Multilevel statistics showed that greater case and death rates (1000 cases or 100 deaths/million inhabitants) in one’s state increased the odds of being fearful of contracting the disease (by 18% and 25%). For each additional 1000 cases or 100 deaths, the odds of currently not working or treating emergencies increased by 36% and 58%. The reduction in patients seen weekly per dentist was greater in public (38.7±18.6) than in private clinics (22.5±17.8). This study provides early evidence of three major impacts of the pandemic on dentistry in Brazil: increasing inequalities due to coverage differences between public and private networks; adoption of new clinical routines, which are associated with an economic burden; and associations of regional COVID-19 incidence and mortality with fear of contracting the disease at work.

## INTRODUCTION

Brazil has emerged as a new COVID-19 pandemic epicenter with steadily growing caseloads. By June of 2020, Brazil was the country with the second-most cases and third-most deaths (COVID-19 Dashboard 2020). With dentistry being a context of high contraction risk and the international supply of personal protective equipment (PPE) compromised, the pandemic has brought major challenges to the dental sector, including maintaining universal dental care coverage for 211 million people dispersed across an 8.5-million-km^2^ area. Brazil, which has more than half a million dental professionals (348,000+ dentists; Federal Council 2020) and accounts for an approximately 2.5% share of the 29+ billion USD global market (360 Reports 2020), has the most important dental industry in Latin America.

While high-quality technological dentistry is available in the private sector, low-income citizens depend on public healthcare systems, which are struggling to cope with the pandemic (Silva et al. 2020). Dentistry personnel are facing new routines, more expensive and less comfortable PPE, fewer appointments, and less revenue. These challenges are superimposed upon already existing economic instability that has persisted since mid-2014. In this context, dentists are challenged with fears of contracting COVID-19 while working in a quickly-changing, turbulent situation that continues to worsen throughout Latin America. Dental teams need to make preventive care efforts to ensure that they do not contribute to worsening the epidemiology of the pandemic. Moreover, the situation is likely to get worse due to Brazil being in a region of developing countries with entrenched inequalities (Rodriguez-Morales et al. 2020).

Planning medium- and long-term actions to respond to the challenges facing the dental sector related to the COVID-19 pandemic will require establishing an understanding of current baseline parameters, including estimates of key resources, of the sector. Accordingly, we conducted the present nationwide survey study in Brazil, the aims of which were to assess COVID-19 pandemic effects on (1) dental coverage, (2) dental office routines and economic burdens, and (3) the behavior of dentists.

## METHODS

### Study design

The study protocol was approved by our institutional research ethics board (#4.015.536). A short questionnaire was developed, pre-tested, and used in a cross-sectional survey with a large sample of dentists in Brazil. In accordance with open science practices, the research protocol, questionnaire in its original language, databank of responses, and other information related to this study are published in an open platform (doi:10.17605/OSF.IO/DNBGS). An English translation of the questionnaire is provided in the Appendix (Table A1). SURGE reporting guideline (Grimshaw, 2014) was consulted. This report does not cover the full survey content. A methodological article detailing how the study was conducted will be published elsewhere.

### Questionnaire development and pre-testing

A self-administered questionnaire about the impact of the pandemic on dental practice routines was developed through consultation with eight researchers in three discrete review rounds. The questionnaire was hosted online (Google Forms). To obtain information about the reliability and validity of the tool and items, we conducted a pre-test in a sample of 22 dentists who were asked to evaluate its clarity, writing style, question sequence, and internal consistency. The pre-testers were asked to respond the questionnaire and record the time to complete; the mean time to complete ± standard deviation (SD) was 7±2 min. Pre-testers scored the clarity of each question on a scale of 1 (not clear) to 5 (very clear). A text box was available after every question for pre-testers to explain their scores and place comments, critiques, suggestions, and other response options. All items with a score ≤3 (n=9) were discussed by at least three researchers to obtain a consensus regarding how to improve them based on pre-tester feedback and then edited accordingly. The mean clarity scores ± SD were 4.79±0.10 for the 9 items that needed revision, and 4.91±0.11 for all 30 items considered together. The questionnaire was reviewed and revised iteratively by the executive group for approval. Pre-testers were precluded from participating in the main study to avoid response bias.

### Questionnaire content

The participant had to click ‘Yes’ after the question “Do you agree to participate in the study voluntarily?” to access the questionnaire. The definitive questionnaire contained 30 mandatory close-ended items, divided into three sections: demographic and professional profile (n=8); professional practices during the pandemic (n=11); and structure and routine of the respondent’s main workplace (n=11). The main outcomes were related to the professionals’ behavior regarding their clinical routines. The options ‘I’d rather not say’, ‘I don’t know how to answer’, and ‘Does not apply’ were available to avoid response errors, and were treated as missing data (see the Appendix for details about questionnaire content).

### Participant recruitment and survey administration

A source population of 24,126 registered dentists who work in the public network (list provided by the Brazilian Ministry of Health) were sent email invitations to participate. The email contained a brief statement that included the study objective, the average response time, notification of the university conducting the study, and a website link to the questionnaire. The initial emails were sent on May 15, 2020; reminder emails were sent 5 days later. Additionally, we created an Instagram® social networking campaign targeting dentists in Brazil (Facebook, Menlo Park, CA). To our best knowledge, this is the first study to use Instagram to recruit healthcare professionals. This social network is highly used by dentists in Brazil; as of, June 11, 2020, there were 5 million and 6.5 million posts with #dentistry and #odontologia (Portuguese for dentistry). The campaign, which started on May 20, invited dentists to participate in an online survey regarding the impact of the pandemic on their practices. An Instagram professional account was created (@odcovid) with a website link to the questionnaire in its bio page. Invitations were posted calling for the participation of dentists; they included the same information provided in the email invites and a hyperlink to the questionnaire was available on the @odcovid bio page. We used hashtags related to dentistry and COVID-19 to increase reach to the target population. Participating researchers shared the invitations on their personal Instagram profiles (feed and stories) and asked other dentists to aid in disseminating the campaign. Brazilian dentists with professional Instagram profiles were asked to also share the invitation post. We reached professionals categorized as micro (<10,000 followers) and meso (10,000–1 million) on the followers scale (Boerman, 2020). A second Instagram campaign with similar content but a slightly different visual presentation was created 2 days later.

### Sample selection and collection of responses

All dentists practicing in Brazil were eligible. Given a target population of ∼348,000 professionals, we estimated that 2,385 responses would be necessary to ensure a 95% confidence interval and 2% margin of error. Responses were collected between May 15 and May 24, 2020.

### Data analysis

Partial questionnaire completion was not possible. In some cases, responses were restricted to a specific population. Descriptive statistics were used to identify frequencies and distributions of variables. Responses to questions on numbers of patients assisted weekly, before and after the pandemic, were subjected to one-way ANOVA. Proportions were compared using chi-square tests. COVID-19 case and death counts in each Brazilian state were obtained from official Ministry of Health reports (Fassa and Tomasi 2020) on May 20, 2020, the date when the greatest number of survey responses was received. For analysis purposes, data were converted into thousands of cases and hundreds of deaths per one million inhabitants in each state. Multilevel mixed effect models were used to test the association between the contextual status of the pandemic in each state and dentistry-related outcomes. Outcomes included decrease in number of patients assisted weekly (numerical), fear of contracting COVID-19 at work (no/a little vs. yes/a lot), and current work status (normal/reduced vs. not working/emergencies only). Linear and logistic models were used for numeric and binary outcomes. The models considered two levels of organization: dentist (level 1) and state (level 2). β-coefficients and Odds Ratios (OR) were reported. Contextual level variance was assessed using intraclass correlation coefficient (ICC) for linear models and Median OR for logistic models (α=0.05). All analyses were performed in Stata 14.2 (StataCorp, College Station, TX).

## RESULTS

A total of 3,122 valid responses were received over 10 days from all 26 Brazilian states and the federal district (gathering of responses over time shown in Fig.1A). The first 5 days included only email invitation responses. The response rate in this period was 2.1%; the numbers of actual rejections/losses cannot be calculated. We received 1,572 responses in the first 24 h after the Instagram campaign started. Respondents were most female (75%) and in practice for ≤20 years (74%). Meanwhile, 53% were working mainly in private clinics, whereas 36% were working in the public sector (Table 1). The mean age ± SD of the respondents was 38 ± 11 years.

**Figure 1.**
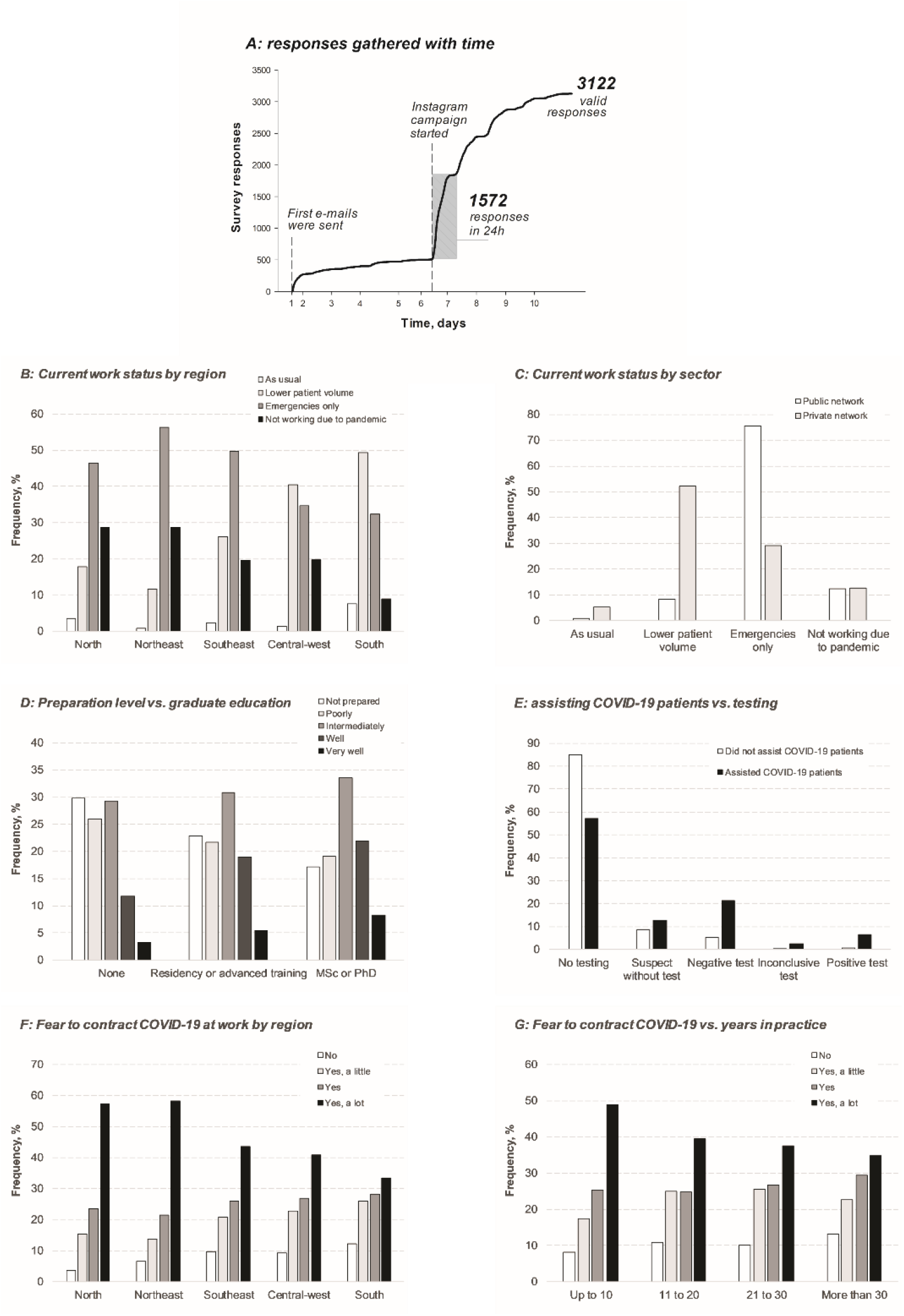
Factors influencing COVID-19 pandemic effects on dental practices. (A) Over 10 days, 3,122 valid survey responses were received from all regions in Brazil. (B) The work statuses of ‘not working’ or ‘emergency only’ were more frequent in the less developed North and Northeast regions (p<0.001). (C) Work status by sector: 52% of private dentists reported seeing less patients than usual, while most public dentists reported emergency appointments only (p<0.001). (D) Education level influenced how prepared professionals feel to assist COVID-19 patients (p<0.001). (E) Dentists who had confirmed contraction of COVID-19 themselves (6.4%) were more likely (p<0.001) to have assisted patients with COVID-19 (tested positive) than dentists who had not (0.7%). (F) Fear of contracting COVID-19 at work varied across regions, being higher in the North and Northeast regions than in other regions (p<0.001). (G) Fear of contracting COVID-19 at work was influenced by years in practice (p<0.001).

**Table 1.** Demographic and work practice characteristics of the respondents, Brazil, 2020 (N=3,122)

| Variable/category | n* | % | 95% CI |
| --- | --- | --- | --- |
| **Gender** | 3,116 |  |  |
| Male | 790 | 25.4 | 23.9; 26.2 |
| Female | 2,326 | 74.7 | 73.1; 76.2 |
| **Years in practice** | 3,121 |  |  |
| ≤10 | 1,496 | 47.9 | 46.2; 49.7 |
| 11–20 | 812 | 26.0 | 24.5; 27.6 |
| 21–30 | 501 | 16.1 | 14.8; 17.4 |
| >30 | 312 | 10.0 | 9.0; 11.1 |
| **Postgraduate education (completed)** | 3,121 |  |  |
| None | 758 | 24.3 | 22.8; 25.8 |
| Residency or advanced special training | 1,530 | 49.0 | 47.3; 50.8 |
| MSc or PhD | 833 | 26.7 | 25.2; 28.3 |
| **Main work sector** | 3,051 |  |  |
| Public | 1,091 | 35.8 | 34.1; 37.5 |
| Private | 1,601 | 52.5 | 50.7; 54.2 |
| Other | 359 | 11.8 | 10.7; 13.0 |
| **Brazilian regional division** | 3,122 |  |  |
| South | 1,183 | 37.8 | 36.2; 39.6 |
| Southeast | 923 | 29.6 | 28.0; 31.2 |
| Central-west | 221 | 7.1 | 6.2; 8.0 |
| Northeast | 682 | 21.9 | 20.4; 23.3 |
| North | 113 | 3.6 | 3.0; 4.3 |
| **Current work status** | 3,056 |  |  |
| As usual | 119 | 3.9 | 3.3; 4.6 |
| Lower patient volume | 994 | 32.5 | 30.9; 34.2 |
| Emergency appointments only | 1,325 | 43.4 | 41.6; 45.1 |
| Not working due to pandemic | 546 | 17.9 | 16.5; 19.3 |
| Not working due to other reasons | 72 | 2.4 | 1.9; 3.0 |
| **Volume of weekly patients compared with pre-pandemic period** | 2,812 |  |  |
| Increased or normal | 62 | 2.2 | 1.7; 2.8 |
| Reduced | 2750 | 97.8 | 97.2; 98.3 |
| **Have you had online patient appointments during the pandemic?** | 2,832 |  |  |
| No but I am willing to do | 755 | 26.7 | 25.1; 28.3 |
| No and I am not willing to do | 1,159 | 40.9 | 39.1; 42.7 |
| Yes, the overall experience was positive | 726 | 25.6 | 24.1; 27.3 |
| Yes, the overall experience was negative | 192 | 6.8 | 5.9; 7.8 |
| **Impact of pandemic in work routine** | 3,048 |  |  |
| No impact | 17 | 0.6 | 0.3; 0.9 |
| Low | 99 | 3.3 | 2.7; 3.9 |
| Intermediate | 389 | 12.8 | 11.6; 14.0 |
| High | 926 | 30.4 | 28.8; 32.0 |
| Very high | 1,617 | 53.1 | 51.3; 54.8 |
| **Have work routine changes led to increased financial costs?** | 2,207 |  |  |
| No | 447 | 20.3 | 18.6; 22.0 |
| Yes, but prices were not adjusted | 1,432 | 64.9 | 62.9; 66.9 |
| Yes, and prices were adjusted for patients | 328 | 14.9 | 13.4; 16.4 |
| **Training for COVID-19 specific preventive measures** | 3,099 |  |  |
| None | 559 | 18.0 | 16.7; 19.4 |
| Online training or general instructions | 2,406 | 77.6 | 76.1; 79.1 |
| Practical training | 134 | 4.3 | 3.7; 5.1 |
| **Have you treated patients with a confirmed COVID-19 diagnosis?** | 2,401 |  |  |
| No or do not know | 2,275 | 94.8 | 93.8; 95.6 |
| Yes | 126 | 5.3 | 4.4; 6.2 |
| **How prepared do you feel to treat patients with COVID-19?** | 3,040 |  |  |
| Not at all prepared | 702 | 23.1 | 21.6; 24.6 |
| Poorly prepared | 670 | 22.0 | 20.6; 23.5 |
| Intermediately | 948 | 31.2 | 29.6; 32.9 |
| Well prepared | 547 | 18.0 | 16.7; 19.4 |
| Very well prepared | 173 | 5.7 | 4.9; 6.6 |
| **Do you fear to contract COVID-19 at work?** | 3,024 |  |  |
| No | 295 | 9.7 | 8.7; 10.9 |
| Yes, a little | 643 | 21.3 | 19.8; 22.8 |
| Yes | 781 | 25.8 | 24.3; 27.4 |
| Yes, a lot | 1,305 | 43.2 | 41.4; 44.9 |
| **Have you suspected or tested yourself for COVID-19?** | 3,093 |  |  |
| No | 2,517 | 81.4 | 80.0; 82.7 |
| Suspect without test | 314 | 10.2 | 9.1; 11.3 |
| Negative test | 213 | 6.7 | 6.0; 7.8 |
| Inconclusive test | 16 | 0.5 | 0.3; 0.8 |
| Positive test | 33 | 1.1 | 0.7; 1.5 |
| **Do you agree with current social distancing measures in your city?** | 3,104 |  |  |
| Fully disagree | 63 | 2.0 | 1.6; 2.6 |
| Partially disagree | 330 | 10.6 | 9.6; 11.8 |
| Not agree or disagree | 38 | 1.2 | 0.8; 1.7 |
| Partially agree | 1,001 | 32.3 | 30.6; 33.9 |
| Fully agree | 1,672 | 53.9 | 52.1; 55.6 |

\*Varies from total N because of missing data for different questions.

### Dental care coverage

Current work status was reported to be affected by 94% of the respondents. Only 2% reported normal or increased patient volumes. Not working/emergency only statuses were more common among dentists working in the less developed North and Northeast regions (Fig.1B). Interestingly, 59% of respondents reported be willing to assist or having already assisted patients online, and 26% regarded such virtual consults as being positive experiences.

Whereas only 52% of private dentists reported seeing less patients than usual due to the pandemic, 76% of public clinic dentists reported maintaining only emergency appointments (Fig.1C), yielding a significant difference on the effect of the pandemic on the volume of patients treated weekly (Table 2). Before the pandemic, the public network covered more patients per dentist. During the pandemic, reductions in weekly dental care levels were reported to be 23 patients/private dentist and 39 patients/dentist in the public network.

**Table 2.** Mean numbers of patients treated weekly per dentist by work sector (standard deviation), before and during the pandemic, Brazil, 2020 (n=2,534 dentists)

|  | Public network | Private practice | Total |
| --- | --- | --- | --- |
| Before pandemic | 47.3 (19.7) | 34.2 (20.8) | 39.6 (21.3) |
| During pandemic | 8.6 (8.6) | 11.7 (13.6) | 10.2 (11.8) |
| Difference* | 38.7 (18.6) | 22.5 (17.8) | 29.2 (19.8) |

\*ANOVA $p < 0.001$ .

The effects of COVID-19 confirmed-case and death rates on the numbers of patients assisted (Table 3) showed dentists seeing two fewer patients/week for each 1000 cases per one million inhabitants, and three fewer patients/week for each 100 deaths. This effect was more pronounced in the public network: 2.45 and 3.25 fewer patients were seen each week for every 1000 cases or 100 deaths per one million inhabitants, respectively. In this analysis, the number of patients seen by dentists working in private practice was not significant.

**Table 3.** Effect of numbers of confirmed COVID-19 cases and deaths * on differences in numbers of patients seen by work sector, Brazil, 2020 (n=2,534 dentists)

|  | Effects on decreases in numbers of patients seen |  |  |  |
| --- | --- | --- | --- | --- |
| | $\beta$ | 95% CI | P-value | ICC |
| **Overall** |  |  |  |  |
| 1000 cases/million inhabitants | 1.96 | 0.43; 3.49 | 0.012 | 0.086 |
| 100 deaths/million inhabitants | 2.90 | 0.80; 5.00 | 0.007 | 0.085 |
| **Public network** |  |  |  |  |
| 1000 cases/million inhabitants | 2.45 | 0.55; 4.36 | 0.012 | 0.144 |
| 100 deaths/million inhabitants | 3.25 | 0.98; 5.52 | 0.005 | 0.137 |
| **Private practice** |  |  |  |  |
| 1000 cases/million inhabitants | 1.12 | -1.55; 3.78 | 0.410 | 0.144 |
| 100 deaths/million inhabitants | 2.34 | -0.95; 5.62 | 0.163 | 0.137 |

\*Multilevel linear regression model considering all 26 different Brazilian states and the federal district. CI, Confidence Interval; ICC, Intraclass Correlation Coefficient.

### Routine and economic burden for dentists

The impact of the pandemic on clinic routines was considered high or very high by 84% of respondents (0.6% reported no impact). Though 80% of respondents reported increased financial costs, only 15% adjusted prices for patients. The pandemic required infrastructural changes in the work setting for 74% of dentists. Most had new types of PPE available for all clinical appointments, including face shields (84%), N95 masks (71%), and disposable coats (66%). Patient screening became more expensive and time consuming due to antimicrobial mouthwashes (46%), completion of COVID-19 questionnaires (35%), and temperature monitoring (24%) mainly. For 35% of respondents, N95 masks were the predominant mask used (with at least half of patients). Taking into account the pre-pandemic volume of patients treated weekly by this sample (average 39.6), and the 348,000 dentists in Brazil, generalizing the figure of 35% of dentists treating at least half of their patients wearing N95 masks without re-use, dentists can be expected to use some 9.6 million masks per month. Considering typical prices for surgical masks (0.4 USD) and N95 masks (2.92 USD) (quotes retrieved by authors), the yearly cost of this simple PPE change would be ∼290 million USD, which would amount to approximately 1.16 billion USD over a potential 4-year COVID-19 resurgence risk period (Kissler et al. 2020).

### Behavior of dentists during pandemic

As reported in Table 1, more than four out of five dentists reported undergoing at least some training in COVID-19 preventive measures, though fewer than one in twenty participated in practical in-clinic training. While almost a quarter of respondents reported feeling well/very well prepared to treat patients with COVID-19, only 5.3% had done so (Table 1). Perception of preparedness to provide care for COVID-19 patients was influenced by education level (Fig.1D). It was more common for dentists who treated patients with COVID-19 to also have COVID-19 (6.4%), than for those who had not seen COVID-19 patients (0.7%) (Fig.1E). Testing was also more frequent for dentists who had seen COVID-19 patients. Although 90% feared contracting the disease at work, only 8% indicated that they had been tested for COVID-19 (1.1% had a positive test). Fear varied among regions, being particularly elevated in the North and Northeast (Fig.1F), and with years in practice (Fig.1G). Fear of contracting COVID-19 at work related positively to the numbers of cases and deaths reported in the state in which the respondent was working. Each 1000 cases per million inhabitants and each 100 deaths per million inhabitants increased the odds of having fear to contract COVID-19 (Table 4). Likewise, MOR indicated that, compared to dentists in less impacted states, dentists practicing in more highly impacted states had a more than 30% greater likelihood of fearing that they may contract COVID-19 and were more than twice as likely to be offering emergency only appointments or to be closed altogether rather than maintaining a usual or even reduced volume of patients with full-service availability. For each 1000 cases and each 100 deaths per million residents in the state, the likelihood of not working or treating emergencies only, as opposed to working with a reduced or typical patient volume, increased by 36% and 58%.

**Table 4.** Effect of numbers of confirmed COVID-19 cases and deaths * on fear of contracting COVID-19 at work and current work status, Brazil, 2020 (n=3,021 dentists)

| Variable | OR | 95% CI | P-value | MOR |
| --- | --- | --- | --- | --- |
| **Fear to contract COVID-19 at work (ref: none/a little)** |  |  |  |  |
| 1000 cases/million inhabitants | 1.18 | 1.01; 1.39 | 0.039 | 1.32 |
| 100 deaths/million inhabitants | 1.25 | 1.02; 1.52 | 0.029 | 1.34 |
| **Work status (not working/only urgencies vs. normal/reduced frequency)** |  |  |  |  |
| 1000 cases/million inhabitants | 1.36 | 1.00; 1.86 | 0.050 | 2.28 |
| 100 deaths/million inhabitants | 1.58 | 1.06; 2.38 | 0.026 | 2.22 |

\*Multilevel logistic regression model considering all 26 different Brazilian states and the federal district. CI, Confidence Interval; OR, Odds Ratio; MOR, Median OR.

## DISCUSSION

Here, we report the findings of the first survey, to the best of our knowledge, in which both email and Instagram social networking campaigns were used to reach healthcare professionals. Although the use of social media in research has been discussed (Schroeder 2014; Ngai et al. 2015; Weller 2015), there is scarce information regarding its use to recruit hard-to-reach populations (Guillori et al. 2018). We felt that a combined strategy was important to recruit dentists working in both public and private networks, and doing so allowed us gather one of the largest samples to date for a COVID-19 survey in the dental field (Ahmed et al. 2020; Cagetti et al. 2020; Consolo et al. 2020; Duruk et al. 2020; Kamate et al. 2020; van der Tas et al. 2020). The pandemic may have facilitated our recruitment owing to people spending more time at home and on social media (Farooq et al. 2020). Although the representativeness of the sample may have limitations, online surveying methods are particularly important during this time when sanitary measures prevent traditional research approaches. The distributions of responses by region, gender, and age were similar to the overall distributions of dentists in Brazil (Morita et al. 2010), except for a slightly higher response rate from females and from Southern Brazil, where the study was originated. It is worth mentioning that the South and Southeast regions, which have similar human development index and per capita income values (IPEA 2016; IBGE 2019), represent the highest-income regions of Brazil. Notwithstanding, our sample variability was supported by the large numbers of responses received.

The present results provide early evidence of three major aspects being at stake in dentistry in the new pandemic epicenter. First, differences in coverage between public and private clinics suggest an intensification of regional and socioeconomic inequalities. Second, although dentists have a similar fear of contracting COVID-19 as other healthcare providers, they report feeling less prepared to assist patients (Zhang et al. 2020). Third, dentists have adopted new routines and incurred increased costs, which eventually will be transferred to patients or paid by the government in public clinics. The scenario is aggravated by disjointed responses from public agencies and the associated lack of an effective coordinated national response to the pandemic (The Lancet 2020).

Dental sector stakeholders seem to be paying diligent attention, with dental councils and sanitary agencies having already released guidance documents. The vast majority of our study respondents (91%) indicated that they are following official regulatory standards in their new routines, and that they, by and large, have made substantial efforts to cope with the new clinical requirements. The low volume of patients currently being seen reflects a prioritization of PPE supplies for healthcare professionals providing medical treatment to COVID-19 patients as well as Ministry of Health directives to provide care for dental emergencies only. Inadequate PPE has been reported to have a negative impact on the mental health of professionals (Simms et al. 2020). Inadequate public healthcare funding also increases the risk of exacerbating historical inequalities. Meanwhile, a prior economic analysis showed that COVID-19 mitigation/suppression measures will cause financial distress to private dental clinics (Schwendicke et al. 2020). One could argue that pandemic-associated increases in the need for medical devices and PPE, and the emerging vaccine industry, should be favorable to business in the biomedical industry. However, in Brazil, this industry accounts for less than 43% of the national consumption production in general biomedical supplies (ABIMO 2020). KaVo, a major dental company worldwide, recently closed its manufacturing facilities in Brazil, which may be an early sign of employment loss in the sector. Government-aided measures to support PPE supply and biomedical industries could be necessary in the long term.

Training in preventive measures and the use of up-to-date screening methods may be appropriate first steps for dentists to feel better prepared to attend to COVID-19 patients. Individual cognizance and knowledge of pertinent information are important factors in healthcare workers feeling confidence in dealing with and overcoming the pandemic (Zhang et al. 2020). Our multi-level analysis shows that mounting COVID-19 case and death counts are affecting dentists’ behavior. The aerosolized cloud in dental offices is a constant reminder of danger. While patient appointment volumes remain far from pre-pandemic levels, our data indicate that Brazilian dentists are open to the incorporation of telehealth programs, which may, despite its associated challenges, be a good strategy for mitigating the impact of the pandemic, while improving preventive actions and reducing unnecessary referrals (Bavaresco et al. 2020). Even after the contagion curve is flattened, we can expect precautionary changes to dental clinic routines and associated stress to persist for years given that dental professionals will continue to be at high risk of exposure, especially in the event of a future resurgence.

It should be noted that our study design does not allow one to establish cause-effect relationships. Notwithstanding, the present study provides important information about early signs of problems in the dental sector during this period of a steeply inclined COVID-19 contagion curve in Brazil. Future studies will be necessary to monitor how dentists are coping with the pandemic. Data from this study may be useful as a baseline relative to future developments and useful in designing interventions. Brazil is a big player in dentistry worldwide, with a particularly predominant role in Latin America. Unfortunately, given its concentrated effects in the public dental care sector, the pandemic appears to be contributing to a deepening of already marked inequalities in oral health within Brazil, and such effects may extend more broadly into Latin America. Actions taken now will affect how Brazilian dentistry is regarded after the pandemic, and whether Brazil will be a good or bad example of dental practices, especially for neighboring countries. Ultimately, the outlook of the dental sector depends on political, professional, and personal actions in this turbulent period during which major aspects are at stake. Constant monitoring of the situation is encouraged over the course of events in the ongoing pandemic.

## Data Availability

Underlying research materials related to this paper can be accessed in OSF: doi:10.17605/OSF.IO/DNBGS

https://osf.io/dnbgs/

## ACKNOWLEDGEMENTS

This study was financed by Fundação de Amparo à Pesquisa do Estado do Rio Grande do Sul (FAPERGS), Brazil (PRONEX 16/2551-0000471-4), Coordenação de Aperfeiçoamento de Pessoal de Nível Superior (CAPES), Brazil (Finance Code 001) and CAPES/PRINT, Brazil (88881.309861/2018-01). The sponsors had no role in study design, collection, analysis or interpretation of data, writing the report, or decision to submit for publication. We thank the Brazilian Ministry of Health for providing the contact of professionals from the public network. We also thank all persons who helped to disseminate the campaign on Instagram and dentists who volunteered to participate. The authors declare no conflicts of interest associated with this manuscript.

## AUTHOR CONTRIBUTIONS

RRM: contributed to conceptualization, methodology, validation, data curation, supervision, project administration, writing, editing and critical review of the manuscript. MBC: contributed to conceptualization, methodology, validation, data curation, formal analysis, writing and critical review. ABQ, AD, JPL: contributed to methodology, validation, data curation and translation, and proofreading of the manuscript. TPC: contributed to conceptualization, methodology, writing and critical review. OPD: contributed to conceptualization, methodology, writing and critical review. MSC: contributed to conceptualization, methodology, validation, writing, and critical review. GSL: contributed to conceptualization, methodology, data curation, writing, and critical review. FFD: contributed to conceptualization, supervision, funding acquisition, writing, and critical review. Order of authors considered relative contribution and gender equality. All authors gave their final approval and agree to be accountable for all aspects of the work.

